# Adaptive deep brain stimulation for gait using a device embedded inertial sensor

**DOI:** 10.64898/2026.08.10.26360135

**Authors:** Ashwini Oswal, Silvia Santoloce, Mayela Zamora, Noelle Jacobsen, Fernando Rodriguez-Plazas, Tao Liu, Bahman Abdi-Sargezeh, Alex Green, Jeanette Brooks, Dominika Kruszynska, Reiko Ashida, Nagaraja Sarangmat, Alan Whone, Timothy Denison

**Affiliations:** MRC Centre of Research Excellence in Restorative Neural Dynamics, University of Oxford, Oxford, UK; Nuffield Department of Clinical Neurosciences, University of Oxford, Oxford, UK; Southmead Hospital, Bristol, UK; Institute of Biomedical Engineering, University of Oxford, Oxford, UK; Amber Therapeutics, Harwell, Oxfordshire, UK; John Radcliffe Hospital, Oxford, UK; School of Psychology and Neuroscience, University of Bristol, Bristol, UK

## Abstract

Recent studies show that pallidal and subthalamic local field potentials (LFPs) encode locomotor state and can guide adaptive deep brain stimulation (DBS) for gait impairment in Parkinson’s disease. Here, in one participant implanted with the Picostim DyNeuMo-2c, we demonstrate a simpler and more direct approach for inferring locomotor state using the device’s onboard accelerometer. Triaxial acceleration was classified independently on each axis to select among preconfigured stimulation programs. Using a cranially mounted digital twin, we characterized inertial signatures across medication and activity states, developed a classifier that distinguished walking from rest while rejecting tremor, and verified the intended stimulation switches during walking. In an exploratory comparison, a gait-adaptive program improved objective gait measures relative to open-loop stimulation optimised for resting tremor. These findings provide a first-in-human demonstration of the feasibility of device-embedded inertial sensing for gait-responsive DBS. They establish a practical framework for further evaluation in larger cohorts.

## Background – Adaptive Methods for Gait Control in DBS

Gait impairment is among the most disabling and treatment resistant features of advanced Parkinson’s disease (PD)^1,2^. Recent studies showed that neural activity recorded from deep brain stimulation (DBS) electrodes in the subthalamic nucleus or globus pallidus encodes locomotor state, and that real-time decoding of these signals can control activity-responsive DBS to improve gait^3–5^. However, these approaches require neural sensing during stimulation, personalised spectral decoders trained across medication states and external processing that is not yet fully integrated into implantable systems^6^. We therefore asked whether locomotor state could instead be detected directly and more simply using an implantable pulse generator’s onboard accelerometer.

Walking is a mechanical event that produces stereotyped, rhythmic accelerations directly measurable by accelerometers already incorporated into many implantable devices. Unlike neural recordings, inertial signals are not contaminated by stimulation artefacts and can be evaluated for robustness across medication states. For the binary classification of walking versus rest, inertial sensing therefore provides a more direct and interpretable measure of behaviour than a neural correlate^7^. As a technology demonstration, we implemented this approach in one participant implanted with a cranially mounted Picostim neurostimulator running DyNeuMo-2c (Dy2c) firmware^8^.

### An embedded inertial control architecture

We implemented gait detection entirely on the device (**Supplementary Figure. 1**). The Picostim DyNeuMo-2c supports both manual and adaptive control of stimulation. In adaptive mode, the embedded triaxial accelerometer samples activity at 3.2 kHz. Each axis is assessed independently, and a classifier recognises common postures together with periods of activity and inactivity. The classifier then selects which preconfigured stimulation program the stimulation engine applies at any moment. A parallel manual pathway allows the adaptive mode to be switched on or off; when it is off, the device reverts to a predefined fallback program. Every transition is logged, and the design records the algorithm state, the active program and the fallback, consistent with IEC 60601-1-10 principles for physiological closed loop controllers^8^.

### Inertial signatures across patient states

To gather the information needed to design the classifier, we monitored the system with a cranium mounted digital twin instrumented to expose all internal device states and sampled triaxial acceleration at 100 Hz while the participant moved through four states: medication OFF while seated, medication OFF while walking, medication ON while seated, and medication ON while walking, with stimulation applied throughout (**Figure. 1**). Gait steps were most prominent on the vertical (z) axis, appearing as punctuated accelerations in both medication states, whereas conducted tremor projected mainly onto the x and y axes, reaching the head through inertial coupling from the arm. Gait and tremor could therefore be separated both by their level and by their axis of orientation, providing a directly interpretable signature of walking that was robust to tremor.

**Figure. 1.**
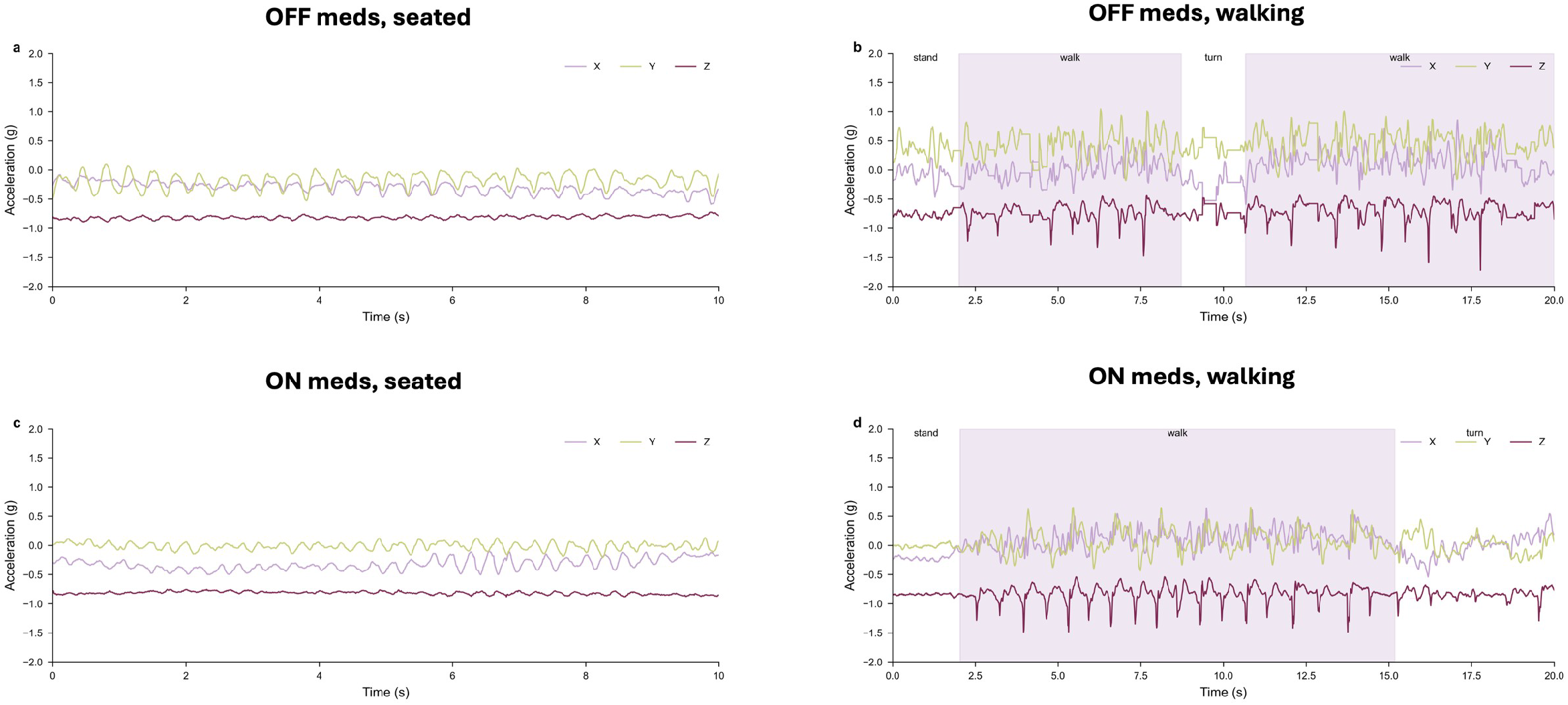
Interpretable inertial signature from a digital twin. Triaxial acceleration sampled at 100 Hz on a cranium mounted digital twin across four states (medication OFF or ON, seated or walking; stimulation applied throughout). Gait steps are most prominent on the vertical (z) axis, while conducted tremor projects mainly onto the x and y axes, reaching the head by inertial coupling from the arm. Gait and tremor are separated by both level and axis orientation.

### A classifier for gait versus rest that drives program switches

From these signatures, we built a simple classifier to distinguish gait from rest while rejecting tremor, using the vertical axis for step detection. On the digital twin, the classifier detected gait onset at the first step and produced the intended control action, switching from the resting program (focus) to the gait program (motion) and reverting to focus once a sustained interval of inactivity was observed (5 s on the twin, extended to 30 s in the participant to tolerate brief pauses in walking) (**Supplementary Figure. 2**). The gait program favoured walking at the cost of some residual breakthrough tremor. These settings were transferred to the participant and confirmed in use, after which the participant was discharged home and was able to walk freely with no external sensors or processing hardware. Program parameters are provided in the Supplementary Methods.

### Gait measures under the gait program versus the tremor program

Finally, we compared gait during the adaptive stimulation program with gait during the open-loop resting program optimised for tremor (Open loop – focus). Gait was quantified with bilateral foot worn Opal inertial sensors and APDM Mobility Lab software, using three interleaved free walking blocks for each condition (**Figure. 2**). In this single participant, median right sided gait speed was higher during the adaptive stimulation program compared to the open loop resting program (0.735 versus 0.660 m/s; median difference 0.075 m/s; FDR adjusted p=0.018), while the smaller increase on the left side was not significant (0.775 versus 0.740 m/s; difference 0.035 m/s; p=0.183). Cadence was higher for the adaptive stimulation program on both sides (left 79.30 versus 71.11 steps/min, difference 8.19, p<0.001; right 78.17 versus 71.31 steps/min, difference 6.86, p<0.001). Median signed stride length asymmetry shifted towards zero (−4.99 versus -13.08 percent; difference 8.09 percentage points; p=0.0013). These comparisons are exploratory (one-sided permutation tests with Benjamini–Hochberg correction) and, in a single case, illustrate a possible benefit of the motion adaptive paradigm on gait rather than establishing clinical efficacy, which will require blinded assessment, longer follow up, and a larger cohort.

**Figure. 2.**
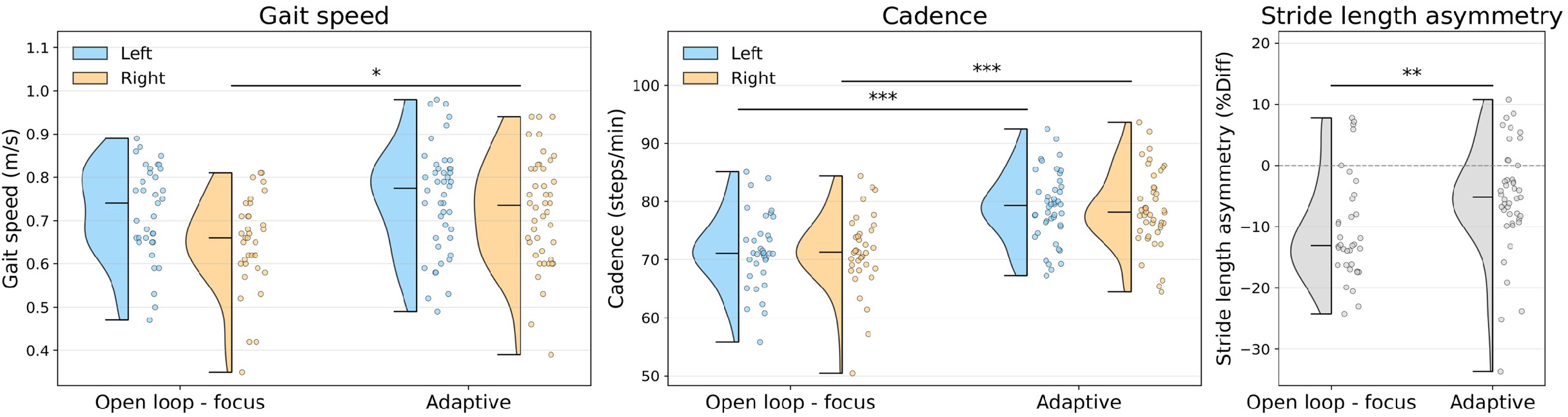
Gait characteristics under open loop focus versus adaptive stimulation. Gait metrics were extracted with APDM Mobility Lab from bilateral foot worn Opal inertial sensors, using three interleaved free walking blocks for each condition. Half violin plots show the distributions of (A) side specific gait speed, (B) cadence, and (C) stride length asymmetry; individual points are stride level observations. Blue and orange indicate left and right sided gait cycles, respectively, and asymmetry is shown in grey. Black markers denote the minimum, median, and maximum of each distribution, with vertical lines spanning the range; the dashed line in C indicates zero asymmetry. Horizontal bars indicate significant exploratory one-sided permutation comparisons testing whether the median was greater during Adaptive than Open loop focus (5,000 label permutations; Benjamini–Hochberg false discovery rate correction across five comparisons; *FDR p<0.05, **p<0.01, ***p<0.001).

## Discussion

We propose motion-adaptive stimulation as a deliberately simple approach, demonstrated here as a technology demonstration. The sensed signal is directly interpretable and tied to what the patient is doing, making it a practical basis for classification. For binary walking-versus-rest detection, a low-cost motion sensor is well matched to the task. As a single case, the gait findings are exploratory, and a blinded study in more patients with longer follow-up is needed before any clinical benefit can be claimed.

Many implanted neurostimulators already contain an accelerometer^7^, so this approach needs no new hardware and mainly motivates activating those sensors as a pragmatic adaptive mode. The power cost is modest. In this device, the accelerometer draws roughly 70 µW against roughly 2.2 mW for bilateral stimulation - about 3% of stimulation power and a smaller proportion of total device power once other functions, such as radio operation and standby currents, are included.

The next generation of commercial accelerometers consume less than 1% of stimulation power, suggesting that the overhead can be small. Prior kinematic adaptive DBS studies have used wearable inertial sensors with external or PC-in-the-loop processing to detect gait changes and modulate stimulation, demonstrating feasibility but relying on an additional hardware-processing chain that a fully embedded system avoids^9–11^.

The same framework may also support more than one motion classifier. With appropriate device configuration, additional states such as tremor can be identified, as suggested by the tremor signature observed in this case, allowing therapy to respond to multiple activity states. Such movement states may be difficult to distinguish reliably from LFPs, providing a further rationale for matching the sensor to the signal. Furthermore, because this approach does not rely on neural sensing, the stimulation programs are not subject to the sensing–stimulation compatibility constraints and interlocks of devices such as the Medtronic Percept^12^ and Newronika AlphaDBS^13^. An additional practical advantage is timescale. Motion sensing is well matched to brief sit-to-stand-to-walk transitions that dominate everyday behaviour, whereas LFP-based adaptation may better capture slower state changes^14^. In this participant, the motion program triggered at the first step and reverted after 30 s of inactivity, supporting adaptation during short walking bouts without external sensors or processing hardware.

## Conclusion

For the specific task of detecting walking and adjusting stimulation, we demonstrated the feasibility of a motion-adaptive system using a low-cost, low-power sensor that provides a directly interpretable measure of behavioural state. Selecting the simplest sensor suited to the control objective may lower the barrier to embedded adaptive stimulation, while reserving neural decoding for applications that require information available only from neural signals.

Further studies are needed to establish reliability and clinical benefit across patients and real-world activities.

## Data Availability

All data produced in the present study are available upon reasonable request to the authors.

## Supplementary figure legends

**Supplementary Figure. 1.**
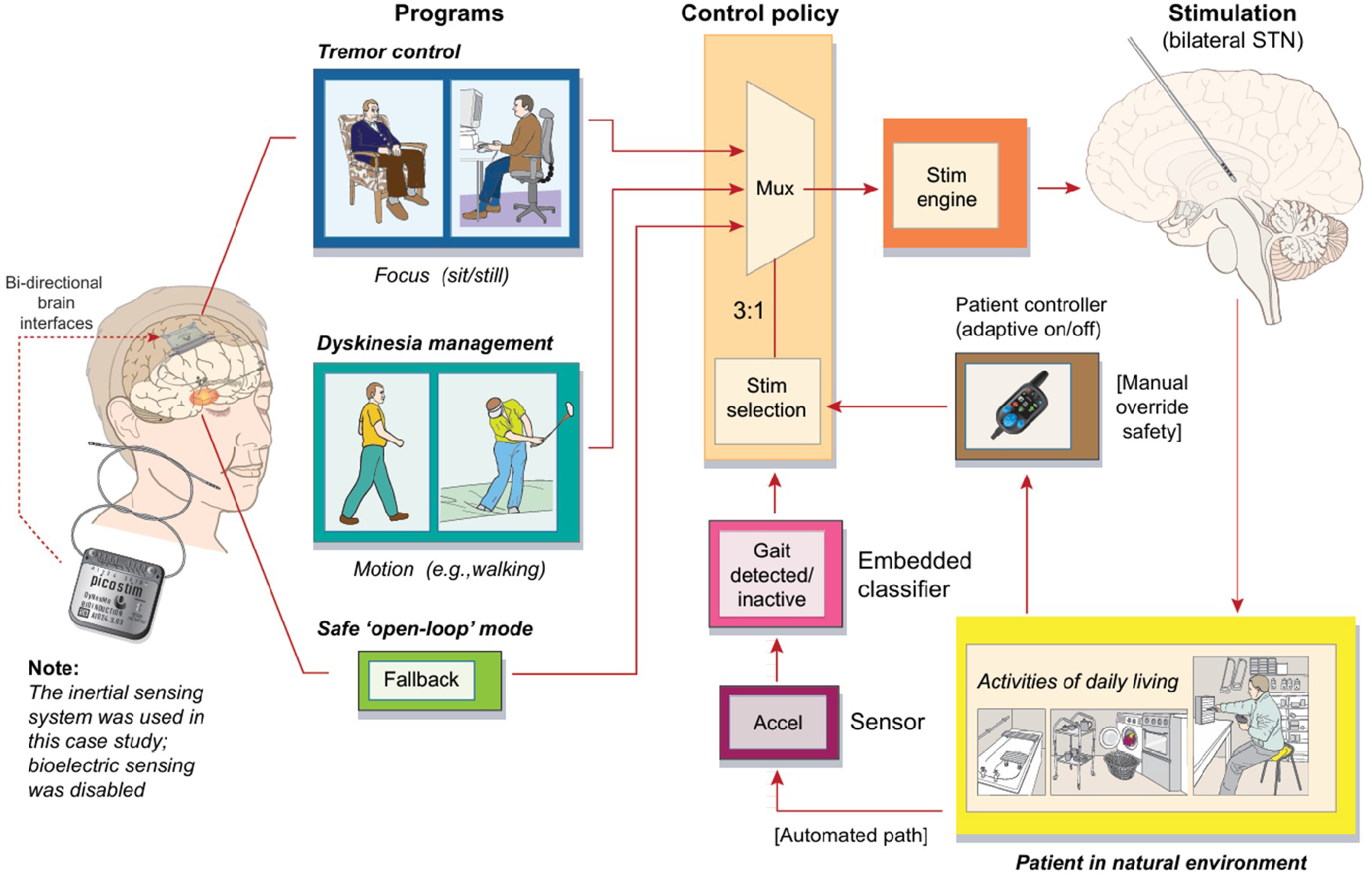
System block diagram. The Picostim DyNeuMo-2c supports manual and adaptive stimulation control. In adaptive mode the embedded triaxial accelerometer is sampled at 3.2 kHz, each axis is assessed and classified independently, and the classifier recognises common postures and periods of activity or inactivity. The classifier selects the preconfigured stimulation program applied by the stimulation engine. A parallel manual pathway turns the adaptive mode on or off; when off, stimulation defaults to the fallback program.

**Supplementary Fig. 2.**
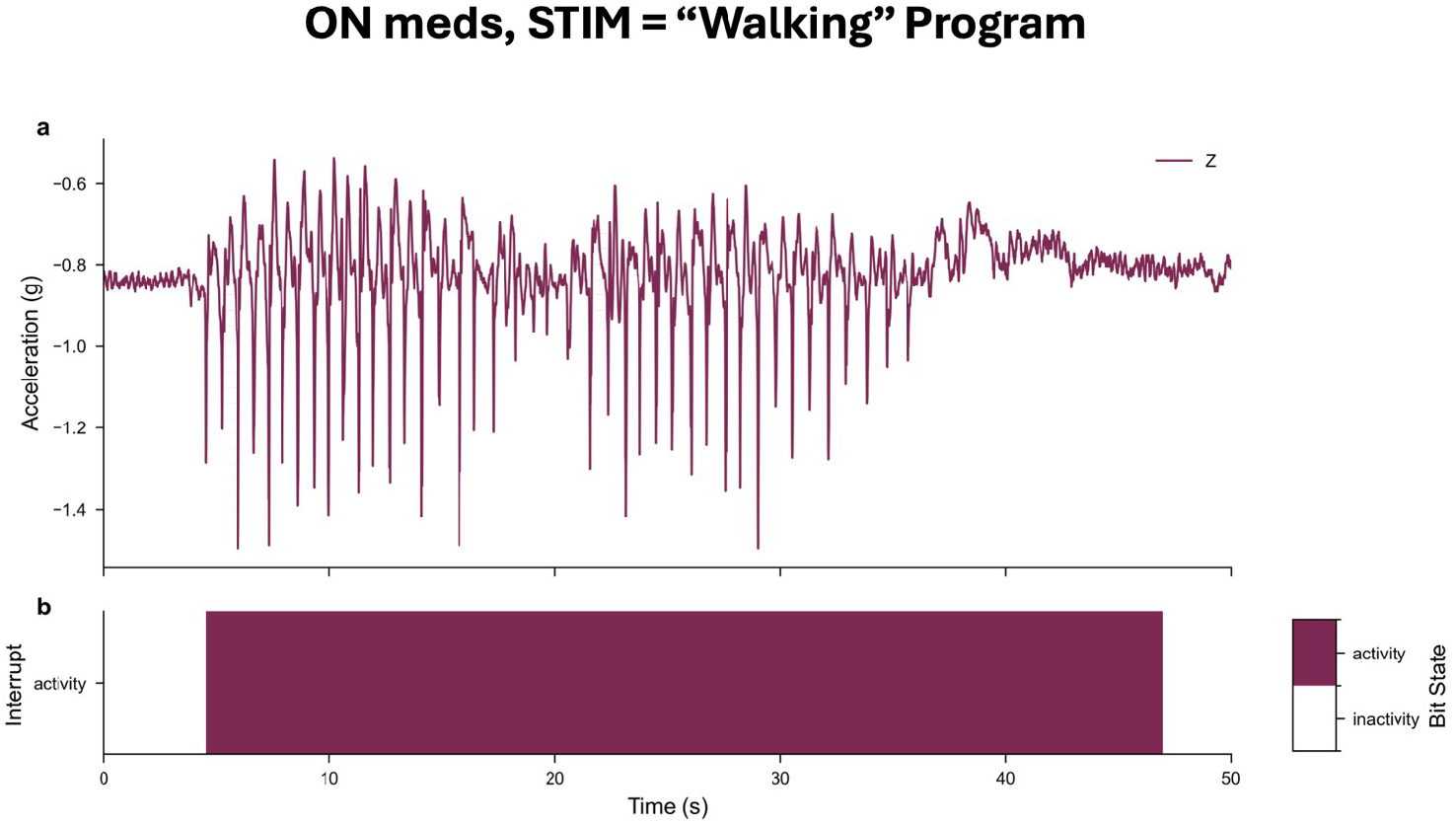
Classifier timing and translation to the patient. Digital twin verification of fully embedded gait onset detection at the first step and transition to the motion program to aid gait at the cost of residual breakthrough tremor, with reversion to the focus program after a sustained inactivity interval (5 s on the twin, 30 s in the participant). Home deployment required no external sensors or processing hardware.

## Author contributions

A.O., A.W. and T.D. conceived and designed the study. M.Z., N.J. and T.D. developed the embedded algorithm and the digital twin. A.O., S.S., N.J., R.A., F.R-P., T.L., B.A-S., N.S., A.G. and A.W. contributed to clinical implementation, programming and data collection. A.O., N.J., and T.D. analysed the data. A.O. and T.D. wrote the manuscript, and all authors reviewed and approved the final version.

## Competing interests

T.D. is a founder and Chief Engineer of Amber Therapeutics, which develops the Picostim and DyNeuMo platforms used in this study. T.D. also serves as a non-executive director of Mint Neuro and ONWARD Medical. A.W. and S.S. have received speaker honoraria from Medtronic. The other authors declare no competing interests.

## Acknowledgements

This work was in part supported by a donation from David Medlock via the Southmead Hospital Charity. A.O. is funded by an MRC Clinician Scientist Fellowship (MR/W024810/1).

## Supplementary Methods

### Participant and regulatory pathway

The participant is a man in his mid-50s with a greater than 10-year history of idiopathic Parkinson’s disease and motor complications refractory to medical therapy. He had been implanted bilaterally with STN DBS leads connected to a Picostim cranially mounted neurostimulator (Amber Therapeutics) as part of the SPARKS first-in-human clinical investigation (IRAS 166307; South West–Central Bristol Research Ethics Committee reference 15/SW/0309; MHRA Clinical Investigation CI/2019/0006; ClinicalTrials.gov identifier NCT03837314). Following the 12-month study period, the participant provided informed consent to continue stimulation under a humanitarian device exemption (HDE), avoiding explantation of a functioning investigational device and interruption of established therapy.

Subsequent deterioration in tremor control, dyskinesia and balance despite optimised open loop reprogramming motivated a further HDE, and additional informed consent and regulatory approval were obtained to permit a non-invasive firmware upgrade enabling DyNeuMo-2c sensing and adaptive stimulation within the existing implanted system. Additional informed consent was obtained for the work reported here.

### Medication

The prescribed dopaminergic regimen was Madopar dispersible 25/100 mg (08:00); co-careldopa 25/100 mg (08:00, 11:00, 14:00, 17:00, 20:00); amantadine 100 mg (08:00, 12:00); rasagiline 1 mg once daily; and co-careldopa CR 100 mg twice nocte. The participant self-reported actual intake at 08:30, 11:30, 13:30 and 16:30 for the short-acting formulations and at 20:30 for the slow-release formulation.

### Digital twin and inertial characterisation

A digital twin of the implant was mounted on the cranium and instrumented to expose all internal device states. The embedded triaxial accelerometer was sampled at 100 Hz while inertial signatures were recorded across four states (medication OFF or ON, seated or walking; stimulation applied throughout). The vertical (z) axis was used for step detection, and conducted tremor was confirmed to project mainly onto the x and y axes. Classifier amplitude thresholds and the inactivity timer were configured on the twin before transfer to the device.

### Stimulation programs

Focus (rest): left lead 80 to 20 current shaping across contacts 1 and 2, 4.5 mA, 70 µs, 125 Hz; right lead contact 6, 3 mA. Motion (gait): left lead redistributed to 50 to 50; right lead was the same as in focus, with a reduced amplitude of 2.6mA.

### Control policy and deployment

The control policy mapped classified inertial state to program selection: gait onset at the first step triggered a transition to motion, and a sustained interval of inactivity triggered reversion to focus (5 s on the twin, 30 s in the participant). Transitions, algorithm state and active program were logged; a fallback program was defined per IEC 60601-1-10. Objective gait was quantified from wearable inertial sensors (APDM Opal) analysed with Mobility Lab, with metrics including gait speed, cadence, and stride length asymmetry. Exploratory one-sided permutation tests (5,000 permutations) with Benjamini Hochberg false discovery rate correction compared adaptive stimulation against open loop focus. The participant was discharged home with settings in place and no external sensors or controllers.

### Statistical note

Single participant proof of concept; data are presented descriptively and any statistical comparisons are exploratory. A multi participant, blinded, crossover trial is required before clinical conclusions can be drawn.

## Notes

### Clinical Trial

NCT03837314

### Author Declarations

The South West Central Bristol Research Ethics Committee (reference 15/SW/0309) gave ethical approval for this work.

## References

1. Bloem, B. R., Okun, M. S. & Klein, C. Parkinson’s disease. The Lancet 397, 2284–2303 (2021).

2. Hausdorff, J. M., Gruendlinger, L., Scollins, L., O’Herron, S. & Tarsy, D. Deep brain stimulation effects on gait variability in Parkinson’s disease. Mov. Disord. 24, 1688–1692 (2009).

3. Louie, K. H. et al. Adaptive deep brain stimulation for dynamic gait control in Parkinson’s disease: a randomized feasibility trial. Nature Medicine 2026 1–12 (2026) doi:10.1038/s41591-026-04434-2.

4. Scafa, S. et al. Activity-dependent adaptive deep brain stimulation improves gait in Parkinson’s disease. Nature Medicine 2026 1–16 (2026) doi:10.1038/s41591-026-04432-4.

5. Balachandar, A. & Fasano, A. Adaptive brain stimulation 2.0 for Parkinsonian gait. Nature Medicine 2026 1–3 (2026) doi:10.1038/s41591-026-04489-1.

6. Zamora, M. et al. DyNeuMo Mk-1: Design and pilot validation of an investigational motion-adaptive neurostimulator with integrated chronotherapy. Exp. Neurol. 351, 113977 (2022).

7. Liu, T. et al. Device-embedded accelerometry complements neural signals for tracking parkinsonian motor states. bioRxiv 2026.07.08.737286 (2026) doi:10.64898/2026.07.08.737286.

8. Benjaber, M. et al. A clinical grade neurostimulation implant for hierarchical control of physiological activity. bioRxiv https://doi.org/10.1101/2025.10.21.683630 (2025) doi:10.1101/2025.10.21.683630.

9. Karjagi, S. et al. Real-Time Kinematic Adaptive Deep Brain Stimulation Safely Reduces Gait Impairment and Freezing of Gait in Parkinson’s Disease. medRxiv https://doi.org/10.64898/2026.02.23.26346487 (2026) doi:10.64898/2026.02.23.26346487.

10. O’Day, J. J. et al. Demonstration of Kinematic-Based Closed-loop Deep Brain Stimulation for Mitigating Freezing of Gait in People with Parkinson’s Disease. Annu. Int. Conf. IEEE Eng. Med. Biol. Soc. 2020, 3612–3616 (2020).

11. Malekmohammadi, M. et al. Kinematic Adaptive Deep Brain Stimulation for Resting Tremor in Parkinson’s Disease. Movement Disorders 31, 426–428 (2016).

12. Sanger, Z. T. et al. Neural signal data collection and analysis of PerceptTM PC BrainSense recordings for thalamic stimulation in epilepsy. J. Neural Eng. 21, 10.1088/1741-2552/ad1dc3 (2024).

13. Arlotti, M. et al. A New Implantable Closed-Loop Clinical Neural Interface: First Application in Parkinson’s Disease. Front. Neurosci. 15, 763235 (2021).

14. Bronte-Stewart, H. M. et al. Long-Term Personalized Adaptive Deep Brain Stimulation in Parkinson Disease: A Nonrandomized Clinical Trial. JAMA Neurol. 82, 1171–1180 (2025).

